# The risk of introducing SARS-CoV-2 to the UK via international travel in August 2020

**DOI:** 10.1101/2020.09.09.20190454

**Authors:** Rachel A. Taylor, Catherine McCarthy, Virag Patel, Ruth Moir, Louise A Kelly, Emma L Snary

## Abstract

International travel poses substantial risks for continued introduction of SARS-CoV-2. As of the 17^th^ August 2020, travellers from 12 of the top 25 countries flying into the UK are required to self-isolate for 14 days. We estimate that 895 (CI: 834–958) infectious travellers arrive in a single week, of which 87% (779, CI: 722–837) originate from countries on the UK quarantine list. We compare alternative measures to the 14 day self-isolation (78.0% effective CI: 74.4–81.6) which could be more feasible long-term. A single RT-PCR taken upon arrival at the airport is 39.6% (CI: 35.2–43.7) effective, or equivalently, it would only detect 2 in 5 infectious passengers. Alternatively, testing four days after arrival is 64.3% (CI: 60.0–68.3) effective whereas a test at the airport plus additional test four days later is 68.9% (CI: 64.9–73.0) effective. Rapidly implementing control measures for travellers from risky countries is vital to protect public health; this methodology can be quickly updated to assess the impact of any further changes to international travel policy or disease occurrence.

## Introduction

In the wake of the SARS-CoV-2 pandemic, causing COVID-19 disease, international air travel effectively stopped [1]. With cases within many countries declining, understanding how to safely re-open international travel and to which countries is of key importance. In particular, it is imperative to determine a risk-based entry strategy, and for that strategy to be flexible in order to quickly react to changing disease prevalence worldwide.

We estimate the weekly number of infectious travellers from 25 countries that have most flights to the UK and assess the impact of self-isolation policies. We also evaluate the effectiveness of alternatives to self-isolation, considering the ability of various health measures on arrival to reduce the potential for onward transmission.

## Risk of introducing SARS-CoV-2

We estimate the weekly number of infectious or pre-infectious travellers with SARS-CoV-2 arriving into each UK airport in an average week in August 2020 (see supplementary material). Infectivity was assumed based on studies reporting serial intervals [e.g. 2] which define a window of infectivity [3]. We assume infectious individuals shed sufficient quantities of the virus to potentially cause a transmission event. Pre-infectious travellers are those that are infected but not yet infectious, and we now refer to both categories of travellers as infectious individuals. We focus on the top 25 countries which fly commercial aircraft to the UK; these countries accounted for 86% of flights into UK airports in August [4]. We estimate expected travel numbers assuming all international travel re-opens but at a lower volume [5]. We assess three types of infectious cases on arrival at the UK airport: 1) a non-UK traveller who is infected in the country prior to travel, 2) a UK traveller returning from the country who was infected prior to travelling to the country, and 3) a UK traveller returning from the country who was infected during their trip. We use estimates of SARS-CoV-2 prevalence from 13^th^ August 2020 in each of the 25 countries, and the UK [6]. Combined in a stochastic assessment that also considers departure detection measures, the travellers decision not to fly due to severe symptoms and the likelihood of being infectious based on their day since exposure, we estimate the expected numbers of travellers on each country-airport route that are infectious on arrival.

The estimated number of infectious cases arriving into UK airports from all 25 countries in a single week in August is 895 (95% CI: 834–958), 0.15% of all expected travellers from these countries. The top 2 countries for import of infectious cases into UK airports, Spain and USA, account for 69.3% of all introductions, with 39.7% from Spain and 29.6% from the USA (Figure 1). Both these countries have large numbers of travellers to the UK normally, and, especially for the USA, prevalence of COVID-19 is high. The USA has the highest rate of infectious travellers per 1000 travellers at 4.58 (CI: 4.08–5.16). Results were most sensitive to uncertainty around flight numbers (Figure S2).

**Figure 1.**
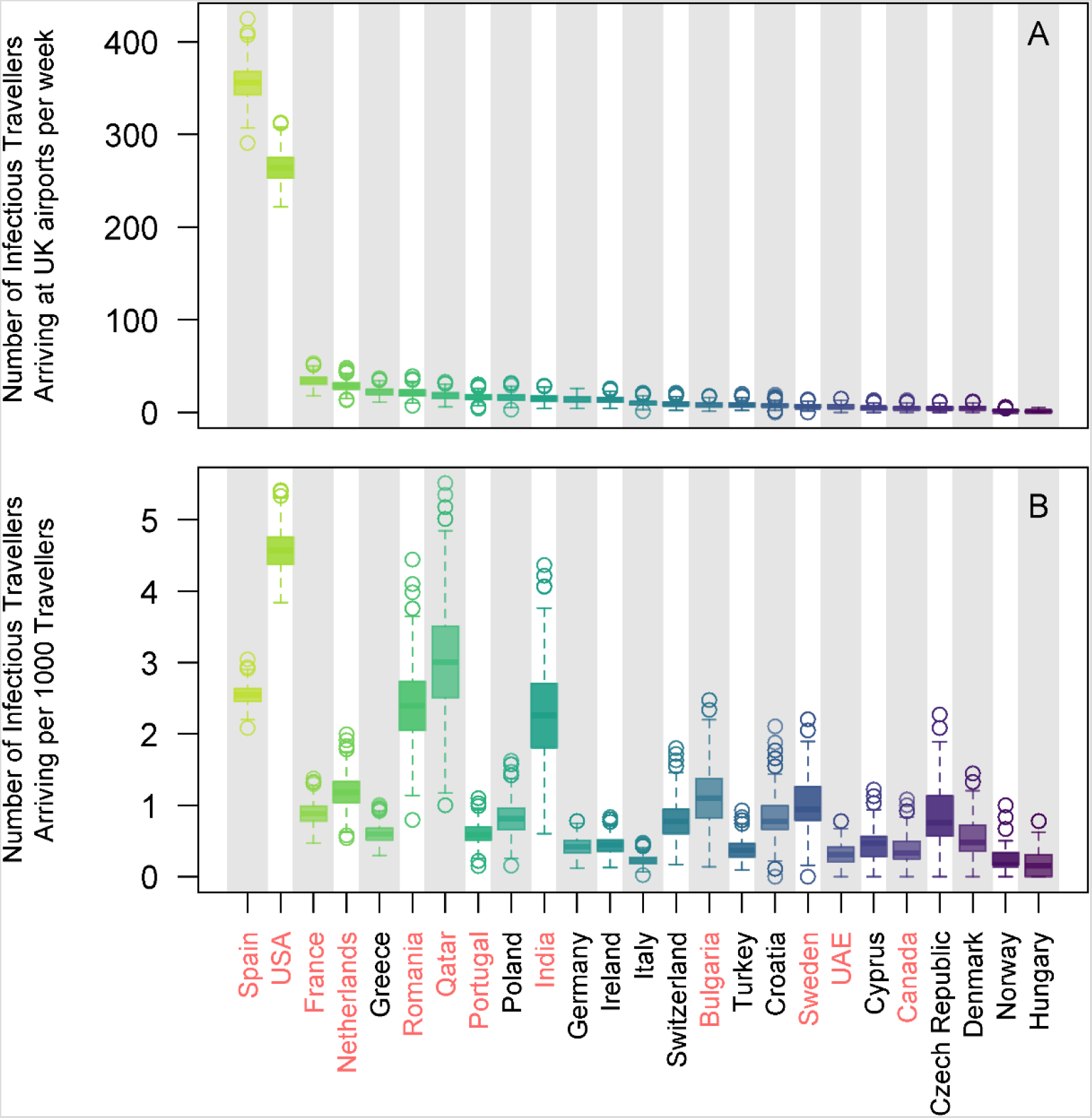
The expected number of SARS-CoV-2 infectious travellers (A), and number per 1000 travellers (B), arriving into UK airports from each of the 25 countries in a single week in August without any UK health measures applied. Countries that require self-isolation on arrival to the UK on 17^th^ August are indicated in red.

The UK is implementing 14 day self-isolation for 12 out of these 25 countries (based on data on 17^th^ August 2020 [7]) with no health measures required for the other 13. The 12 countries with travel restrictions account for 779 (CI: 722–837) of the 895 infectious travellers on arrival (Figure 2A), therefore potentially 87% of infectious arrivals are prevented from onward transmission (depending on efficacy of self-isolation) due to these travel restrictions, with 116 infectious travellers arriving without requiring self-isolation.

Eight of the 12 countries that have travel restrictions are in the top 12 countries, when ranking by the number of infectious travellers (Figure 1A). Notably, Greece has the 5^th^ highest number of infectious travellers, but does not have travel restrictions. Although prevalence in Greece is lower than many other countries [6], it is a popular tourist destination, and hence the expected number of infectious travellers is higher. If the travel restrictions were, instead, to be based on the top 12 countries by number of infectious travellers, 91% (819, CI: 761–876) of all infectious arrivals would need to self-isolate (Figure 2A). Under this strategy an extra 76,334 travellers would require self-isolation (Figure 2B) thereby it has a lower benefit (number infectious self-isolating)–cost (total self-isolating) ratio (BCR). A strategy based on top 12 countries by rate of entry (Figure 1B) has the highest BCR (Figure 2C).

**Figure 2.**
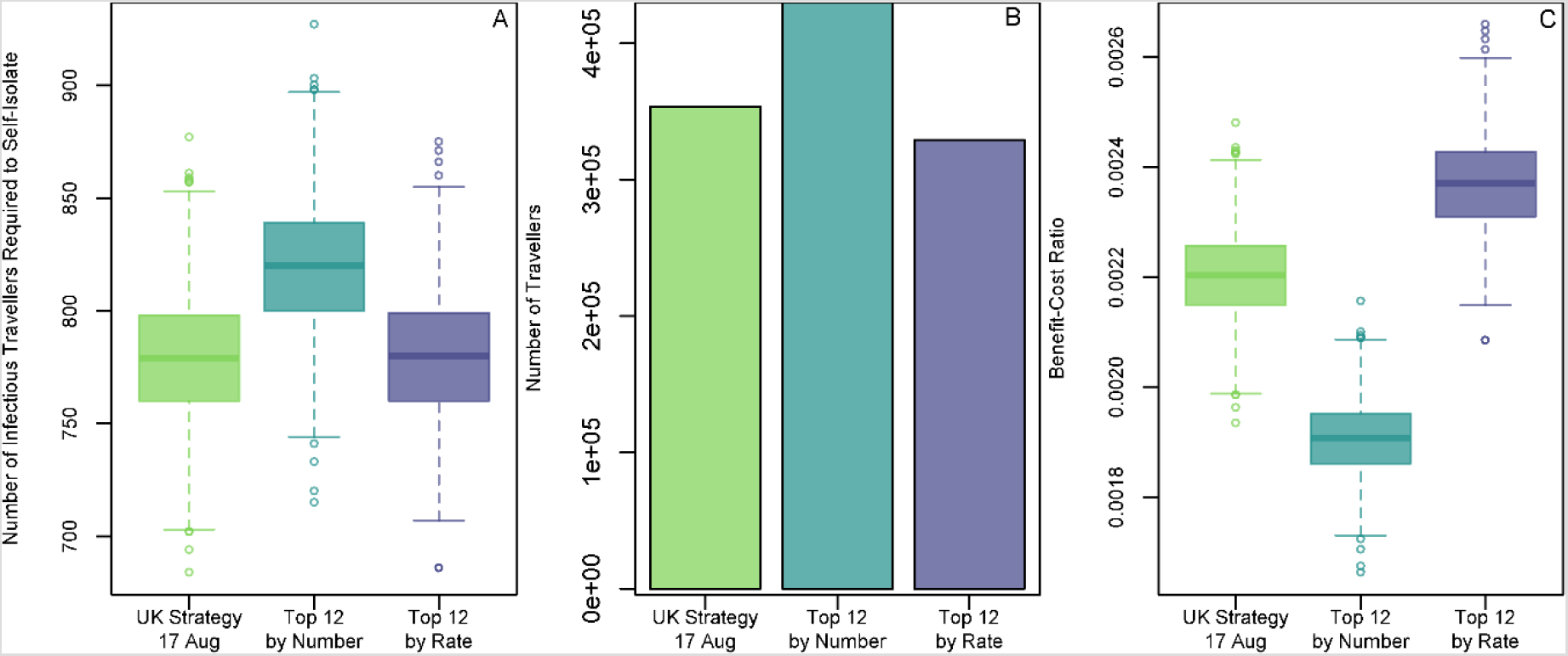
The number of infectious travellers required to self-isolate (A), the total number of travellers required to self-isolate (B) and the BCR (C) for three different travel restriction strategies. The UK strategy is based upon the 12 countries within our 25 that require self-isolation on arrival. “Top 12 by Number” and “Top 12 by Rate” strategies are assuming the top 12 countries by number or rate (respectively) of infectious arrivals, as assessed in Figure 1, require self-isolation on arrival.

## Alternatives to self-isolation

We compare alternative measures to the 14 day self-isolation which could be more feasible long-term. Efficacy is measured by the ability of each health measure to minimise the risk from infectious travellers (either through self-isolation or identifying infectious travellers). We simulate a population of 1 million infected individuals to estimate the probability of showing symptoms or being infectious depending on the day since exposure (see supplementary material). We consider health checks and thermal imaging scanners at the airport, self-isolation upon arrival (for 7, 10 or 14 days) and reverse-transcriptase polymerase chain reaction (RT-PCR) tests under different regimes. For RT-PCR tests, travellers would have to self-isolate until test results are returned (assumed 48 hour delay). We do not consider any ongoing transmission from a traveller who has obtained a positive RT-PCR. We assume a non-compliance rate of 20% for travellers waiting for RT-PCR results, self-isolation periods without RT-PCR and health checks (therefore we do not assume that 14 day self-isolation periods are 100% effective).

Both thermal imaging scanners (0.78% effective, CI: 0.19–1.64) and health checks (1.13%, CI: 0.38–2.09) are completely ineffective at identifying infectious passengers (Figure 3). Airports which are relying on thermal imaging scanners as their only detection method will only identify one in every 128 infectious passengers. A single RT-PCR taken upon arrival at the airport is 39.6% (CI: 35.2–43.7) effective, or equivalently, it would only detect 2 in 5 infectious passengers. The low efficacy is surprising but is due to the high likelihood that a passenger will be pre-infectious at the airport. Alternatively, testing four days after arrival (ST4) is 64.3% (CI: 60.0–68.3) effective whereas a test at the airport plus additional test four days later (DT4) is 68.9% (CI: 64.9–73.0) effective. Given the 48 hour delay in receiving test results, a DT4 regime requires travellers to self-isolate for 6 days. This strategy is statistically superior (by t-test) to self-isolation for 7 days (51.3% effective, CI: 47.2–55.7, p< 0.001), is comparable to self-isolation for 10 days (68.8%, CI: 65.1–72.9, p = 0.93) and less effective than self-isolation for 14 days (78.0% effective CI: 74.4–81.6, p< 0.001). Delaying the RTPCR for 7 days is more effective (ST7: 74.3% effective, CI: 70.0–78.0; DT7: 75.9% effective, CI:72.3–79.6) but requires lengthier self-isolation.

**Figure 3.**
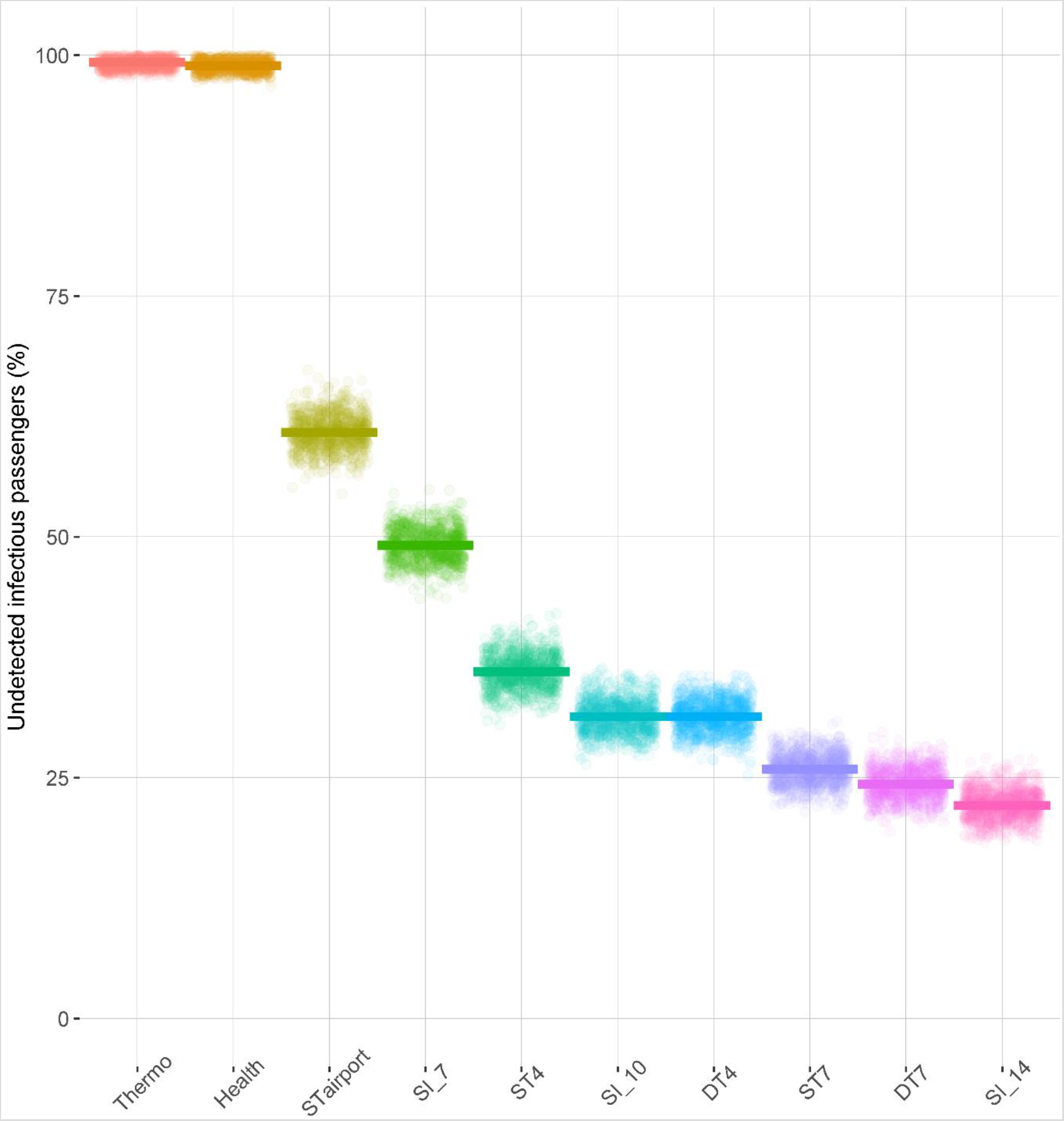
Comparison of SARS-CoV-2 health measures for international travellers. Points show the percentage of infectious passengers which weren’t detected by the health measure on each model iteration. Horizontal bars show mean values. Health measures are thermal imaging scanner (Thermo), health checks (Health), single RT-PCR taken at the airport (STairport), self-isolation for 7 days (SI_7), single RT-PCR taken 4 days after arrival (ST4), self-isolation for 10 days(SI_10), double testing RT-PCR first at the airport and then 4 days after arrival (DT4), single RT-PCR taken 7 days after arrival (ST7), double testing RT-PCR first at the airport and then 7 days later (DT7) and self-isolation for 14 days (SI_14).

## Discussion and conclusions

We show that the current 14 day self-isolation requirements are vital for controlling the risk of continued introduction of SARS-CoV-2 into the UK. The 12 countries from the top 25 which required self-isolation would otherwise be responsible for 779 infectious travellers arriving each week. Only 116 infectious travellers are predicted to arrive from the remaining 13 countries. Even fewer infectious travellers would bypass self-isolation by alternative strategies for quarantine (Figure 2). We use a detailed model of local prevalence and numbers of flights, and show that the three highest risk countries, as of the 13^th^ August 2020 are Spain, the USA and France.

We show that neither thermal imaging scanners, health checks nor a single RT-PCR test at the airport are effective alternatives to self-isolation. Thermal scanners were more ineffective than previously thought [8], but this is in agreement with the lack of efficacy during the Ebola epidemic [9]. The high level of false negatives from these three measures is concerning given that 52% of the top 25 countries are using these methods for departure screening [10]. Individuals who receive a negative RT-PCR result may be less inclined to follow social distancing practices due to a perceived safety around an individual’s personal role in transmission and requirement to return to public-facing workplaces [11]. RT-PCR at the airport and four days after arrival (with a 48 hour wait for results) is 69% effective (compared to 78% for 14 day self-isolation). A double testing strategy is likely to cause financial and logistical challenges [12], but the shorter self-isolation requirements may lead to higher levels of compliance. Lengthy self-isolation periods may also incur indirect costs due to lost workplace productivity and less social expenditure. It is likely that a strategy other than the 14 days self-isolation will be needed for longer term planning.

Further work is required to understand the risk of viral transmission during transit, as has been reported on a flight from the Central African Republic to France [13] and to extend this work to other entry methods, such as ferry or euro-tunnel. We show that the UK guidance is in line with appropriate international risk levels and prevents a significant number of infectious travellers arriving. However, when also factoring in efficacy of self-isolation, this still allows entry of 289 infectious individuals on average per week. It is important that public health officials act rapidly with the latest data estimates in order to control the risk from this fluctuating situation; this methodology can be quickly updated to assess the impact of any further changes to international travel policy or disease occurrence.

## Data Availability

Not applicable

## Acknowledgements

We would like to thank Matthew Coleman (APHA) for review of the model and code and Zach deVries (APHA) for collating data and administrative support. Furthermore, we would like to thank two anonymous reviewers (organised through the Royal Society’s Rapid Assistance in Modelling the Pandemic (RAMP) group) for reviewing the model.

## Author Contributions

ES and RM conceived the study. RT and CM designed the model framework, wrote the model and produced results. VP provided the prevalence estimates. All authors reviewed the model design. RT and CM wrote the manuscript. All authors reviewed the manuscript and gave approval for publication.

